# Exploring the association of subnational drowning mortality and environmental exposures: A global analysis using satellite-derived data

**DOI:** 10.64898/2026.04.19.26351234

**Authors:** Ryan Essex, Samsung Lim, Jagnoor Jagnoor

## Abstract

**Introduction:** Drowning risk begins with water exposure, yet population-water relationships have rarely been quantified at scale using environmental measures. This study explored whether satellite-derived data was associated with subnational drowning mortality and whether associations differed by country income level.

**Methods:** We linked Global Burden of Disease (GBD 2021) age-standardised drowning mortality rates to satellite-derived exposures for 212 subnational regions across 12 countries (2006–2021; 3,392 region-years). Exposures were extracted via Google Earth Engine and standardised. Gamma-log generalised linear mixed models included region random intercepts and year fixed effects. Income-stratified models were estimated separately. Supplementary models assessed maritime vessel activity.

**Results:** Near-water population percentage was the strongest correlate of drowning (IRR 1.40; 95% CI 1.33–1.47). Permanent water coverage was protective (IRR 0.80; 0.73–0.88), as were nighttime lights (IRR 0.96; 0.95–0.97) and hot days ≥30°C (IRR 0.95; 0.92–0.99). Mean temperature (IRR 1.17; 1.11–1.23) and precipitation (IRR 1.03; 1.01–1.04) were positively associated. Near-water effects were consistent across income strata (LIC 1.25; MIC 1.31; HIC 1.24), while other predictors showed weak or inconsistent within-strata associations. Vessel activity was modestly associated with drowning in Global Fishing Watch models (IRR 1.05; 1.01–1.09) but not in Synthetic Aperture Radar models.

**Discussion:** Satellite-derived indicators can characterise drowning risk at scale, with population proximity to water emerging as a robust cross-context correlate. Protective associations for permanent water suggest landscape configuration may shape risk beyond proximity alone, highlighting geospatial data’s value for targeting prevention where surveillance is limited.

## Introduction

Drowning risk starts with exposure to water, whether in or around the home, permanent or seasonal, recreational or accidental. Drowning risk is also shaped by a range of environmental factors. While these statements might seem obvious, drowning prevention efforts have not yet quantified the relationship between populations, water exposure, and other environmental factors influencing risk. Looking toward the periphery (and beyond) of the drowning prevention literature, a small body of work has begun leveraging geospatial and satellite data to understand risk. Satellite imagery has been used to identify populations exposed to flooding and track these changes over time (1) and to map coastal erosion (2). Yet, as directly applied to drowning, this field remains nascent; a 2016 review of geospatial analysis for unintentional injury identified only four drowning-related studies (3). Amongst these studies, the influence of neighbourhood, socioeconomic and built environmental factors were found to be influential in explaining drowning risk (4, 5). Within public health circles more broadly, the potential of such data is being increasingly realised and utilised with such data being used to examine issues air pollution (6) and water quality (7) and in disease surveillance (8). This study seeks to examine how these techniques could be applied to drowning prevention, utilising satellite and geospatial data to explore their relationship to drowning rates, across a sample of sub-national regions, linking satellite data to Global Burden of Disease (GBD) drowning data.

## Aims

This study sought to explore the potential of satellite-derived environmental data in predicting sub-national drowning rates. Specifically, this study had two aims, first to quantify associations between environmental factors and drowning mortality. That is, how variables such as surface water coverage, population proximity to water, built environment characteristics, temperature, and maritime activity were related to age-standardised drowning mortality rates across subnational regions from 2006 to 2021. Second, to examine income-level modification of these associations. That is, whether the relationships between environmental exposure and drowning mortality varied systematically by national income level, in low, middle, and high-income countries.

## Methods

### Overview

This study employed a longitudinal ecological design that. Drowning mortality estimates from the GBD study were merged with environmental exposure data extracted from multiple satellite-based datasets using Google Earth Engine (GEE). Analysis was restricted to subnational regions (ADM1^1^ level) where GBD data provided drowning mortality estimates and where matching to administrative boundaries was possible. Subnational, as opposed to national, regions were selected, as environmental conditions relevant to drowning (e.g. surface water, shoreline complexity, flood-prone population and vessel activity) vary substantially within countries. Aggregating to national level would risk obscuring this heterogeneity, diluting potential associations.

The study proceeded in four stages: (1) creation of environmental exposure and drowning data, which involved extracting environmental exposure variables from satellite datasets for each region-year and matching these to GBD mortality data; (2) descriptive analysis of environmental exposures and drowning rates; (3) statistical modelling to quantify associations and test for income-level effect modification and 4) sensitivity, robustness and model diagnostic checks. Analysis was conducted using GEE accessed through R Studio (9) (version 4.5.0) via the reticulate package (10). A list of all packages used is included in supplementary material (Table S1).

### Creation of environmental exposure data

Data related to environmental exposures came from multiple satellite-based earth observation datasets. All data extraction was performed using GEE (11). For each region-year (2006-2021) a function was utilised in GEE that 1) retrieved region boundaries; 2) created a 50km buffer around the boundary, 3) simplified complex geometries to 5km tolerance (to reduce computational overhead); (4) created water occurrence masks (identifying pixels which were water and those which were not) and distance rasters from global surface water data (see below); and (5) extracted or calculated all environmental variables, including near-water population percentage (population within 500m of water / total population × 100), water coverage percentage (surface water area / total land area × 100), permanent water percentage (permanent water area / total land area × 100), and population density metrics (population / built-up area, both overall and restricted to built-up areas within 500m of water). Table 1 provides a description of each data source, variable (extracted and derived), and temporal aggregation method.

**Table 1:** Environmental data sources and processing methods.

| Data Source | Variable(s) | Temporal Coverage | Spatial Resolution | Temporal Aggregation Method | Description |
| --- | --- | --- | --- | --- | --- |
| **JRC Global Surface Water v1.4 (14)** | Water occurrence, permanent water, shoreline length, shoreline density | 1984-2021 (composite) | Native ~30 m, aggregated to 100 m | Static long-term composite; no temporal aggregation (single occurrence/seasonality products used for all study years). | JRC Global Surface Water maps the presence of surface water, reporting the percentage of time water was detected (0–100%) and the number of wet months (0–12). “Any water” was defined as pixels with >10% occurrence; “permanent water” as pixels with water in all 12 months. Total water area, permanent water area, shoreline length, shoreline density, and a 500 m near-water zone were calculated for each region. |
| **WorldPop 100m Population (15)** | Total population, near-water population | 2000-2020 (annual) | 100m | For each year, we obtained all available map tiles, creating a mosaic for each region, summing total pixel values. | WorldPop provides gridded estimates of population by redistributing census counts using geospatial covariates (e.g., buildings, land cover) at 100 m resolution. For each region-year we summed all population pixels to obtain total population, then applied the 500 m near-water buffer to derive population living close to water. |
| **Global Human Settlement Layer (GHSL) P2023A (16)** | Percentage of built-up surface area, population density in built areas | 5-yearly from 1975 onwards | Native 10 m, aggregated to 100 m | Five-yearly snapshots; for each study year we used the closest available range (e.g., 2010 utilised 2008–2012). | GHSL identifies built structures and estimates built surface area. We summed built-up area per region and within the 500 m near-water zone, calculating the percentage of built up area per region, and a crowding measure (people per km<sup>2</sup> of built surface). |
| **FABDEM (Forest-And-Building-removed DEM) (17)** | Percentage of population in flood-prone areas | Static (2021) | Native 30 m, aggregated to 100 m | Static elevation model; no temporal aggregation | FABDEM is a 30 m global digital elevation model with forests and buildings removed to approximate bare-earth elevation. We identified pixels with elevation <5 m and within 500 m of surface water, then summed WorldPop population within these zones to estimate the percentage of the population exposed to low-lying, flood-prone areas. |
| **ECMWF ERA5-Land (18)** | Mean annual temperature, days exceeding 30°C, wind speed (95th percentile) | 1950-present (hourly) | ~9 km grid | For each region we utilised the 12:00 (noon) time slice for each day, and then calculated mean temperature across the year, counted days with noon temperature ≥30 °C, and the 95th percentile of wind speed. | ERA5-Land provides hourly, land-only atmospheric and surface weather and climate data. For each region-year, we summarised temperature and wind based on midday conditions to characterise mean warmth, frequency of hot days, and atypical high wind speeds. |
| **CHIRPS (Climate Hazards Group InfraRed Precipitation with Station data) (19)** | Annual precipitation | 1981-present (daily) | ~5.5km | Daily rainfall summed within each calendar year to produce annual total precipitation averaged across each region | CHIRPS combines satellite and gauge observations to provide global daily rainfall. For each region-year, we summed daily precipitation to obtain total annual rainfall (mm). |
| **NOAA DMSP-OLS Nighttime Lights (stable lights product) (20)** | Nighttime lights | 1992-2013 (annual composites) | ~1km | We used the single annual “stable lights” composite per year (pre-processed to remove ephemeral lights), mosaicked tiles where needed, and computed mean radiance per region. | DMSP-OLS stable-lights composites capture persistent artificial lighting from settlements and infrastructure, widely used as a proxy for urbanisation and economic activity. For pre-2014 years, these annual composites were used to derive a region-level night-time lights indicator. |
| **NOAA VIIRS Day/Night Band monthly stray-light corrected (21)** | Nighttime lights | 2014-present (monthly composites) | ~500m | Monthly composite images were averaged across all 12 months in each year to produce an annual mean radiance image, then mean radiance was calculated per region. | VIIRS Day/Night Band provides monthly composites of average night-time radiance. Following the end of DMSP-OLS coverage, we used VIIRS to derive annual mean night-time lights from 2014 onwards. |
| **Global Fishing Watch (GFW) global vessel hours (22)** | Vessel hours per km<sup>2</sup>, total vessel hours | 2012-2016 | ~1km | Native: monthly. We selected up to 12 images (one per month where available), summed these to create an annual vessel-hours image, converted to hours per km<sup>2</sup> and 1) calculated mean vessel hours per km<sup>2</sup> and 2) summed vessel hours within each region | Global Fishing Watch aggregates tracked vessel activity into a global grid. Using the all-vessels layer, we applied a monthly sampling strategy (maximum 12 images per year) to reduce computational load while retaining seasonal information. The resulting indicators describe the intensity of vessel activity (hours per km<sup>2</sup>) and total vessel hours for each coastal region-year. |
| **Sentinel-1 SAR GRD (23)** | Total vessels detected, small vessels, vessel density per 1,000 km<sup>2</sup> | 2014-2021 | ~10m | Native: irregular intervals (days to weeks). We selected up to 12 images (one per month where available), counting total vessels, small vessels and vessel density | Sentinel-1 images the ocean surface. For each year, we used up to 12 images over each region (about one per month) and applied a fixed brightness threshold over water to flag likely vessels. We then counted total and small vessels and calculated vessel density per 1,000 km<sup>2</sup> |

Subnational drowning mortality rates were extracted from the Global Burden of Disease Study 2021 (12). Specifically, age-standardised mortality rates (per 100,000 population) for the ‘drowning’ cause category, for both sexes combined, annually from 2006 to 2021 were utilised.

To examine effect modification by economic development, countries were classified into income categories based on World Bank 2021 classifications: low-income countries (LIC), middle-income countries (MIC), and high-income countries (HIC). A time-invariant classification applied to all years in the study period was used. The final sample included two LICs (Ethiopia, Kenya; 9 regions; n=144), six MICs (Brazil, India, Iran, Mexico, Pakistan, South Africa; 109 regions; n=1,744), and four HICs (Italy, Japan, Norway, United States; 94 regions; n=1,504).

To join data, the Food and Agriculture Organization’s Global Administrative Unit Layers (GAUL) 2015 Level 1 (ADM1) boundaries (13) was utilised. These boundaries generally represent the primary subnational administrative divisions within countries, states, provinces, or equivalent jurisdictions. This resulted in 212 matched subnational regions yielding 3,392 region-year observations. Some regions could not be matched due to differences in naming, or because of the fact that boundaries had changed over the period of time in question.

### Analytic strategy

#### Descriptive analysis

After compiling the above dataset, we explored trends and distributions of environmental exposures and drowning rates. For each variable, we calculated means, standard deviations, medians, interquartile ranges, and ranges across all region-years. We also examined temporal trends in drowning rates.

#### Variable selection for modelling

As preliminary analyses revealed substantial correlations between a number of variables, and to be transparent in subsequent modelling, we sought to first identify the most influential variables in relation to drowning mortality. Our candidate predictor set included 14 variables available across the full study period (2006-2021): near-water population percentage, water coverage, permanent water coverage, shoreline density, percentage of population in low-lying flood zones, percentage of built-up area, population density near water, nighttime lights, mean temperature, hot days ≥30°C, high (95th percentile) wind speed, and annual precipitation. We applied elastic net regularization (α=0.5, balancing L1 and L2 penalties) with 10-fold cross-validation to identify predictors most strongly associated with log-transformed drowning rates. We selected predictors with non-zero coefficients at λ = λ.1se to favour parsimony. We compared elastic net results with pure Least Absolute Shrinkage and Selection Operator (LASSO) (α=1.0) to assess selection stability. We assessed collinearity using variance inflation factors (VIFs). Our final predictor set comprised variables that were both found to be influential, which had acceptable collinearity properties, and which were conceptually distinct. This approach led to the selection of seven predictors: near-water population percentage, permanent water coverage percentage, mean temperature, hot days ≥30°C, high (95th percentile) wind speed, annual precipitation and nighttime lights. These findings are summarised as supplementary material in Table S2.

#### Modelling environmental exposure and drowning risk

To facilitate interpretation and comparison of effect sizes, we standardised all continuous predictors to have mean 0 and standard deviation 1 using the full dataset’s means and standard deviations. Our primary models used generalized linear mixed models (GLMMs) with a Gamma distribution and log link, appropriate for continuous outcomes (like mortality rates) that are right-skewed and bounded at zero.

We included random intercepts for regions (n=212) to account for unmeasured time-invariant characteristics (e.g., cultural factors, healthcare system features, water safety infrastructure, for example) and to properly account for within-region correlation across years. We included year as a fixed effect to control for trends in drowning mortality that might reflect changes in prevention efforts, surveillance, for example. Our model took the form:

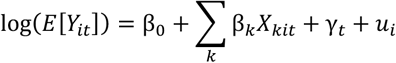

where *Y*_*it*_ was the drowning mortality rate (per 100,000) for region i in year t, *X*_*kit*_ represented standardised environmental predictor k, γ_*t*_ was a year fixed effects, and 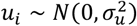 were region-level random effects.

To assess the robustness of associations and understand confounding patterns, we fit three nested models with an increasing number of controls:

- Model 1 (water exposure): percentage near-water population and percentage permanent water coverage, plus year fixed effects and region random effects.
- Model 2 (+ socioeconomic/built environment): Model 1 plus nighttime lights.
- Model 3 (full environmental model): Model 2 plus climate variables (mean temperature, hot days ≥30°C, 95th percentile wind speed, and annual precipitation)

Models were compared using Akaike Information Criterion (AIC) and Bayesian Information Criterion (BIC) to compare fit.

#### Income stratified models

To examine whether associations between environmental factors and drowning differed by national income level, we fit separate models within each income group (low, middle, and high-income countries), using all seven predictors identified through variable selection. This stratified approach allowed all model parameters to vary freely between income groups, revealing context-specific associations without imposing constraints from a pooled model. We compared effect estimates across income groups to identify patterns of effect modification, focusing particularly on water exposure variables (near-water population, permanent water coverage, flood-zone population) and socioeconomic factors (nighttime lights, population density near water) where differential effects by development level were hypothesised. Each income-stratified model retained the same mixed-effects structure (random intercepts for regions, year fixed effects) as the main pooled model.

#### Vessel density models

Because maritime vessel density data was only available for limited temporal windows, we fit separate supplementary models restricted to these periods. Global Fishing Watch (GFW) vessel hour data were available for 2012-2016 (5 years, 1,040 region-year observations), while Synthetic Aperture Radar (SAR) vessel detection data were available for 2014-2021 (8 years, 1,664 region-year observations). These models included the same core environmental predictors as the main model plus vessel density metrics (GFW: vessel hours per km^2^; SAR: small vessel density per 1,000 km^2^, focusing on vessels <20m that may represent subsistence fishing and recreational craft).

#### Sensitivity, robustness and model diagnostics

To assess the stability of our findings, we conducted six sensitivity analyses: 1) Alternative water metrics. We compared models using permanent water coverage versus total water coverage to ensure results were not sensitive to this variable choice; 2) Leave-one-country-out. We systematically excluded each country and refit the main model, calculating the coefficient of variation (CV) for key effect estimates across these leave-one-out samples. We interpreted CV <10% as indicating robust, stable associations; 3) Temporal stability. We fit the main model separately for early (2006-2013) and late (2014-2021) periods to assess whether associations changed over time; 4) Random slopes. We tested whether near-water population effects varied across countries by fitting a model with random slopes for this predictor by country, comparing fit to the random-intercept-only model; 5) Alternative specification. We compared the Gamma-distributed rate model to a negative binomial count model with population offset to ensure findings were not due to outcome distribution assumptions; 6) Spatial clustering. We examined standardised residuals aggregated by country to assess whether residuals showed systematic spatial patterning that might indicate unmeasured confounding or model misspecification.

In terms of model diagnostics, we assessed collinearity among predictors using variance inflation factors. We evaluated overdispersion using likelihood-ratio tests comparing fitted models to saturated models. We inspected diagnostic plots (observed vs. fitted values, residuals vs. fitted, Q-Q plots) to assess model assumptions. For the main model, we identified potentially influential observations using standardised residuals >3 and evaluated model stability after their exclusion.

## Results

### Descriptive Results

Our analytic sample comprised 3,392 region-year observations from 212 subnational administrative regions across 12 countries spanning 2006–2021. Mean drowning mortality rate was 2.63 per 100,000 (SD 1.85). Near-water population percentage averaged 6.56% (SD 5.48) Permanent water coverage averaged 1.45% of regional land area (SD 1.89). Drowning rates remained substantially higher in low-income settings (mean 4.68 per 100,000, n=144 observations from 9 regions in 2 countries) and middle-income settings (mean 3.37 per 100,000, n=1,744 observations from 109 regions in 6 countries) compared to high-income settings (mean 1.58 per 100,000, n=1,504 observations from 94 regions in 4 countries). These findings and a full summary of descriptive statistics are summarised as supplementary material in Tables S3, S4, S5 and Figures S1.

### Main models

The main modelling strategy involved fitting three nested models with an increasing number of controls. Model 1 (water exposure only: near-water population, permanent water coverage) included 2 predictors with 3,392 observations (AIC = 116.6, BIC = 239.2). Model 2 added a socioeconomic proxy (nighttime lights) with 3 predictors and 3,392 observations, improving fit substantially (AIC = 24.3, BIC = 153.0, ΔAIC = −92.3). Model 3 (full environmental model) incorporated 7 predictors with 3,328 observations and further improved fit (AIC = 9.4, BIC = 162.1, ΔAIC = −14.9 relative to Model 2). The substantial improvement from Model 1 to Model 2 (ΔAIC = −92.3) indicated that adding socioeconomic context (nighttime lights) provided major explanatory value beyond water exposure alone, while the further improvement from Model 2 to Model 3 (ΔAIC = −14.9) suggested that climate variables added additional predictive power. These findings are summarised in Table S6.

In the final model (with all variables included), near-water population exposure showed the strongest association with drowning mortality (IRR = 1.40, 95% CI 1.33–1.47, p < .001), suggesting that a one standard deviation increase in near-water population percentage (≈5.48 percentage points) was associated with a 40% increase in drowning rates, after adjusting for other environmental factors. Mean temperature also showed a positive association (IRR = 1.17, 95% CI 1.11–1.23, p < .001), and annual precipitation showed a modest positive association (IRR = 1.03, 95% CI 1.01–1.04, p < .001). Several factors showed protective associations: nighttime lights were protective (IRR = 0.96, 95% CI 0.95–0.97, p < .001), permanent water coverage also showed a protective association (IRR = 0.80, 95% CI 0.73–0.88, p < .001), and hot days ≥30°C showed a modest protective effect (IRR = 0.95, 95% CI 0.92–0.99, p = 0.005). Wind speed (95th percentile) showed no significant association (IRR = 1.01, 95% CI 0.98– 1.04, p = 0.48). These findings are summarised in Figure 1 and Table S7.

**Figure 1.**
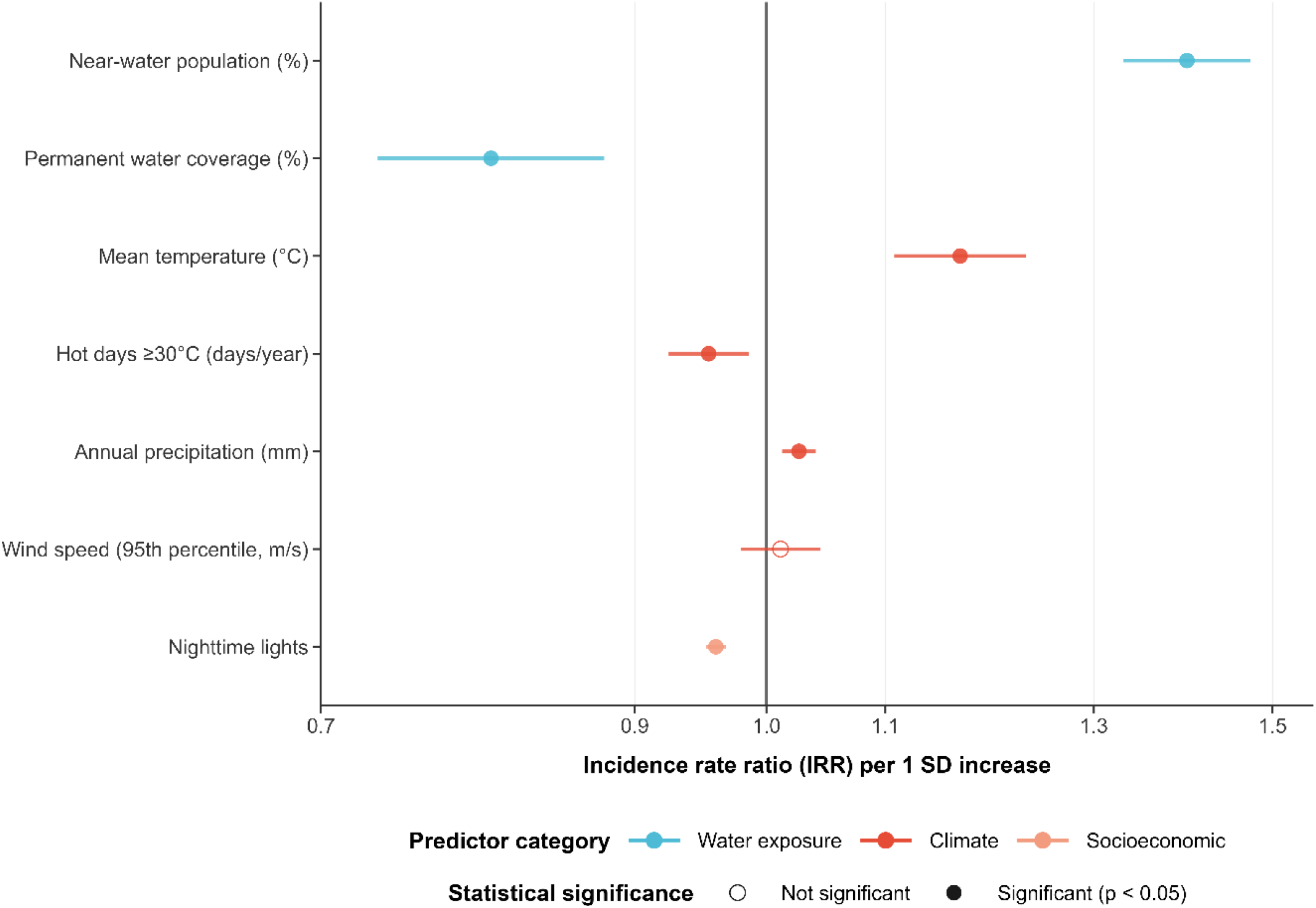
Main model: Predictors of drowning mortality.

### Income stratification

When stratified by income classification (low, middle and high income countries), the relationship between environmental predictors and drowning mortality revealed important patterns across development contexts. Near-water population exposure emerged as a universal risk factor across all income levels. In low-income countries (n=144 observations), each standard deviation increase in near-water population was associated with a 25% increase in drowning mortality (IRR = 1.25, 95% CI 1.09–1.43, p = 0.001). Middle-income countries (n=1,744 observations) showed a 31% increase per standard deviation (IRR = 1.31, 95% CI 1.21–1.42, p < 0.001), while high-income countries (n=1,504 observations) demonstrated a 24% increase (IRR = 1.24, 95% CI 1.18–1.29, p < 0.001). The consistency of this effect across income groups suggests that proximity to water bodies poses drowning risk regardless of economic development level.

Beyond near-water population exposure, few predictors showed clear within–income-group associations. Permanent water coverage was not significantly associated with drowning in any income group (LIC: IRR = 0.94, 95% CI 0.80–1.11, p = 0.49; MIC: IRR = 0.98, 95% CI 0.89– 1.07, p = 0.60; HIC: IRR = 1.02, 95% CI 0.90–1.15, p = 0.77). Nighttime lights similarly showed no association in low-income (IRR = 0.99, 95% CI 0.98–1.01, p = 0.32) or middle-income settings (IRR = 1.01, 95% CI 1.00–1.02, p = 0.10), but remained positively associated with drowning in high-income countries (IRR = 1.01, 95% CI 1.01–1.02, p < 0.001), consistent with the possibility that higher radiance or brightness in more affluent settings is a proxy for greater exposure to recreational or urban water hazards.

Temperature variables did not show significant income stratification. While mean annual temperature was a significant predictor in the overall model, income-stratified analyses revealed no significant temperature associations in any income group (LIC: IRR = 1.14, 95% CI 0.96–1.34, p = 0.13; MIC: IRR = 0.97, 95% CI 0.90–1.04, p = 0.35; HIC: IRR = 1.01, 95% CI 0.97–1.04, p = 0.73). Similarly, hot days ≥30°C, annual precipitation, and wind speed (95th percentile) were not statistically significant within any income group (all p > 0.05), though precipitation showed borderline positive associations in middle-and high-income settings (MIC p = 0.09; HIC p = 0.08). These findings are summarised in Figure 2 and Table S8.

**Figure 2.**
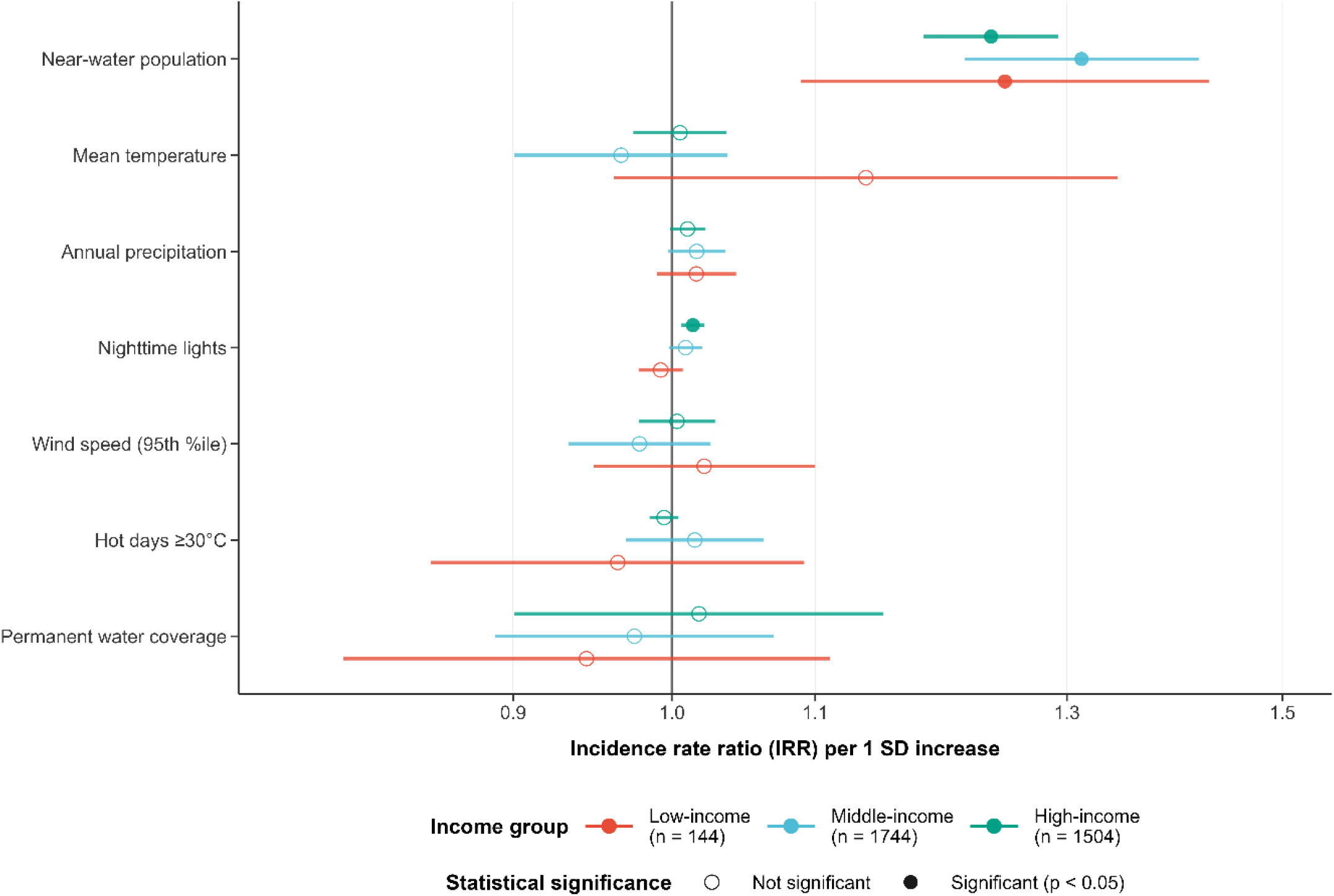
Income stratified covariates

### Vessel models

Among 1,040 region-year observations with Global Fishing Watch (GFW) industrial vessel data (2012–2016), vessel activity showed a modest but statistically significant positive association with drowning mortality (IRR = 1.05, 95% CI 1.01–1.09, p = 0.02). A one standard deviation increase in vessel hours per km^2^ was associated with a 5% increase in drowning rates, controlling for water exposure and other environmental covariates. The near-water population effect remained robust in the GFW sample (IRR = 1.32, 95% CI 1.23–1.42, p < 0.001). Permanent water coverage remained protective (IRR = 0.83, 95% CI 0.76–0.91, p < 0.001), and mean temperature showed a positive association (IRR = 1.21, 95% CI 1.15–1.27, p < 0.001). Annual precipitation was borderline positive (IRR = 1.02, 95% CI 1.00–1.03, p = 0.05), while wind speed, nighttime lights, and hot days ≥30°C were not significant.

Among 1,664 region-year observations with SAR vessel detection data (2014–2021), small vessel density (vessels <20 m) showed no significant association with drowning mortality (IRR = 1.01, 95% CI 1.00–1.02, p = 0.16). The near-water population effect again remained robust (IRR = 1.35, 95% CI 1.26–1.46, p < 0.001), as did the protective effect of permanent water coverage (IRR = 0.84, 95% CI 0.76–0.93, p < 0.001). Mean temperature remained positively associated (IRR = 1.18, 95% CI 1.12–1.25, p < 0.001), while nighttime lights (IRR = 0.95, 95% CI 0.90–0.99, p = 0.01) and hot days ≥30°C (IRR = 0.96, 95% CI 0.93–0.99, p = 0.02) were protective in this subsample. These findings are summarised in Figure 3 and Table S9.

**Figure 3.**
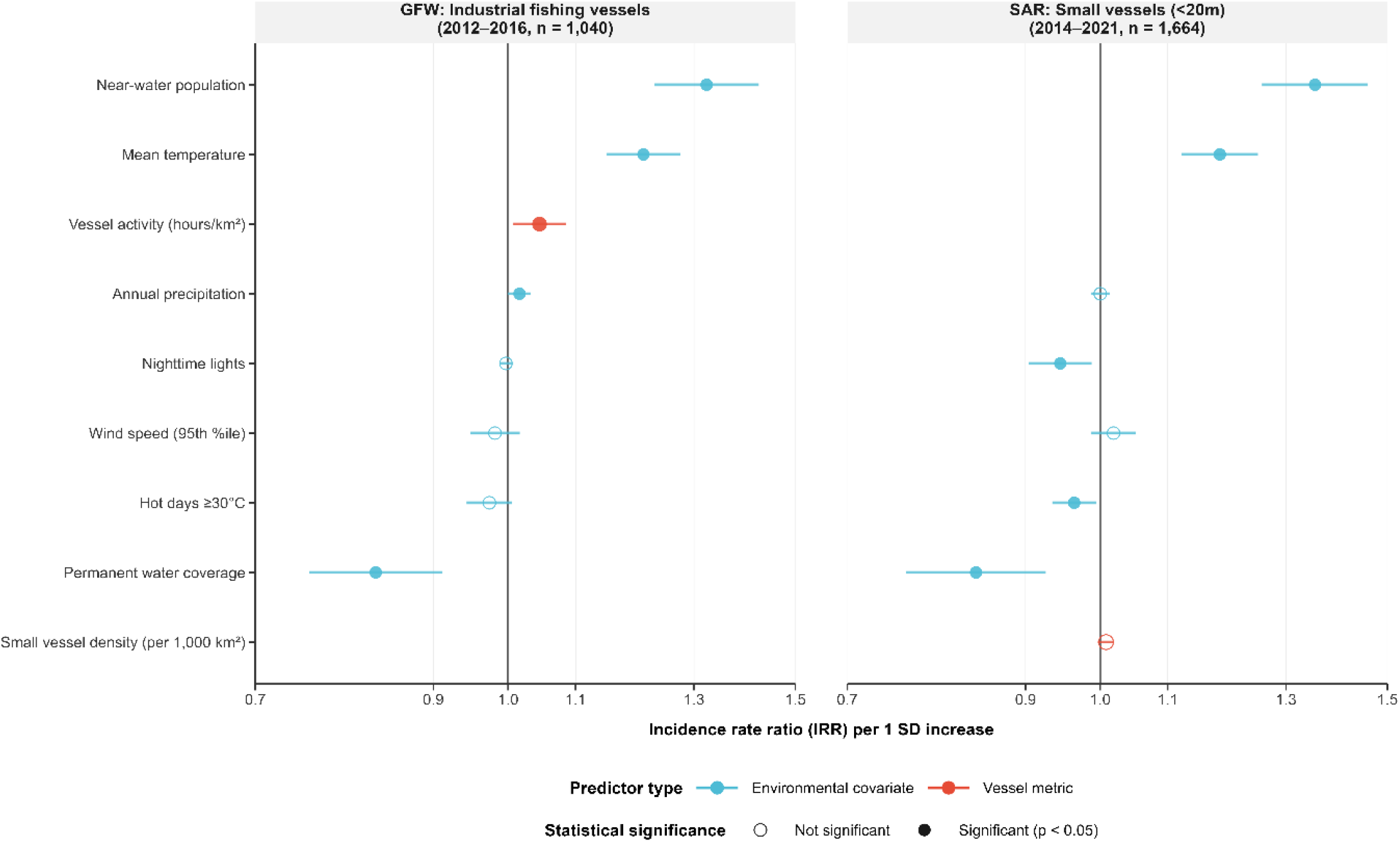
Association between vessel activity and drowning mortality.

### Sensitivity, robustness and diagnostics

Across sensitivity and robustness checks, the overall pattern of associations was consistent: the near-water population measure remained the dominant risk correlate, and key inferences were robust to temporal splitting (Table S10), country influence checks (Tables S11–A12), alternative water-coverage specifications (Table S13), and alternative model forms (Table S14). In the early versus late period comparison, the near-water population effect attenuated only modestly (IRR 1.42 in 2006–2013 vs 1.34 in 2014–2021), while temperature associations were similarly stable (IRR 1.12 vs 1.19), suggesting headline relationships were not driven by a particular sub-period. Country influence analyses indicated no single country drove findings, with individual exclusions changing the near-water IRR by only a few percentage points. Results were also robust to how water coverage was operationalised: substituting “any water coverage” for “permanent water coverage” yielded similarly protective associations while leaving near-water effects unchanged. Alternative model specifications (random slopes, count models, identity-link Gaussian, omitting year fixed effects) did not improve fit relative to the gamma-log specification and generally fit substantially worse. Diagnostics supported model adequacy: collinearity was low (maximum VIF 2.37) with no evidence of overdispersion (dispersion ratio 0.624) (Table S15). Residual screening identified 29 extreme observations (|standardised residual| > 3), but excluding these did not overturn conclusions and strengthened overall fit—consistent with results not being driven by atypical region-years.

## Discussion

This study explored whether satellite-derived environmental data could predict subnational drowning rates and whether associations varied by national income level. Our findings demonstrate that environmental exposures explain meaningful variation in drowning rates, with near-water population exposure emerging as the most consistent risk factor across all contexts (IRR 1.40 pooled; 1.24–1.31 across income strata). Notably, permanent water coverage showed a protective association when controlling for population proximity, suggesting stable water bodies may pose less risk than transient or seasonal water. Income stratification revealed that while near-water effects were universal, other environmental associations were inconsistent within strata, suggesting relationships beyond proximity may be context-specific or driven by between-country differences.

These findings align with broader drowning literature recognising that low- and middle-income countries face disproportionate burden and may require distinct prevention approaches rather than mirroring high-income interventions (24, 25). The inconsistency of other predictors within income groups likely reflects heterogeneity in geography, settlement patterns, and water-use practices, alongside unmeasured social and institutional mediators; supervision norms, water competence, occupational exposures, not captured in these models. The protective effect of permanent water coverage may reflect adaptation; populations living around stable water bodies may develop protective behaviours, infrastructure, or preparedness that transient flooding does not permit (26, 27). Finally, the fact that the number of hot days was protective (i.e. more hot days were associated with lower drowning rates), while mean temperature was a risk factor suggests that associations between drowning and temperature are likely more complicated than simply taking into account mean temperatures. These findings warrant further exploration in future studies.

Several limitations warrant acknowledgment. Our ecological design precludes individual-level inference. GBD estimates incorporate modelling assumptions, particularly where registration systems are limited. Regional averages may mask within-region heterogeneity, and our sample was restricted to 12 countries with available subnational estimates.

This study demonstrates satellite-derived data can meaningfully characterise drowning risk at scale. The global consistency and accessibility of such data offers particular promise for low- and middle-income countries where burden is highest but surveillance limited. Integration of geospatial data could identify high-risk regions, monitor changing risk landscapes, and evaluate whether prevention reaches exposed populations. Drowning is preventable; the tools to map its environmental determinants are available and as this study shows, it is possible to begin to apply this at scale.

## Data Availability

All data used in this study is publicly available

## Footnotes

1 This generally refers to the first-level administrative division of a country, which corresponds to a state, province, or district, depending on the country

